# Impact of COVID-19 Restrictions on People with Hypertension

**DOI:** 10.1101/2020.10.12.20211722

**Authors:** Carissa Bonner, Erin Cvejic, Julie Ayre, Jennifer Isautier, Christopher Semsarian, Brooke Nickel, Carys Batcup, Kristen Pickles, Rachael Dodd, Samuel Cornell, Tessa Copp, Kirsten McCaffery

**Affiliations:** Sydney Health Literacy Lab, School of Public Health, Faculty of Medicine and Health The University of Sydney; Agnes Ginges Centre for Molecular Cardiology at Centenary Institute, The University of Sydney, Australia; Faculty of Medicine and Health, The University of Sydney, Australia; Department of Cardiology, Royal Prince Alfred Hospital, Sydney, Australia

**Keywords:** Covid-19, Hypertension, Anxiety, Case-control

## Abstract

**Objectives:** It is unclear how people with hypertension are responding to the COVID-19 pandemic given their increased risk, and whether targeted public health strategies are needed.

**Design:** This retrospective case-control study compared people with hypertension to matched healthy controls during COVID-19 lockdown, to determine whether they have higher risk perceptions, anxiety and prevention intentions.

**Methods:** Baseline data from a national survey were collected in April 2020 during COVID-19 lockdown. Of 4362 baseline participants, 466 people reported hypertension with no other chronic conditions, and were randomly matched to healthy controls with similar age, gender, education and health literacy. A subset (n=1369) was followed-up at 2 months after restrictions eased, including 147 participants with hypertension only. Risk perceptions, prevention intentions and anxiety were measured.

**Results:** At baseline, perceived seriousness was high for both hypertension and control groups. The hypertension group had higher anxiety than controls; and were more willing to have the influenza vaccine. At follow-up, these differences were no longer present in the longitudinal sub-sample. Perceived seriousness and anxiety had decreased, but vaccine intentions for both influenza and COVID-19 remained high (>80%).

**Conclusions:** Anxiety was above normal levels during the COVID-19 lockdown. This was higher in the hypertension group, who also had higher vaccination intentions. Locations with prolonged restrictions may require targeted mental health screening for vulnerable groups. Despite a decrease in perceived risk and anxiety after 2 months of lockdown restrictions, vaccination intentions for both influenza and COVID-19 remained high, which is encouraging for future prevention of COVID-19.

## INTRODUCTION

Although research on COVID-19 risk factors is constantly evolving, there is consistent evidence that people with cardiovascular disease (CVD) are more likely to experience severe complications and are more likely to die if they acquire COVID-19 (Zaman et al., 2020). People with CVD are more likely to have co-morbidities that may complicate their response to COVID-19, and COVID-19 causes cardiovascular damage (Li, Hu, & Gu, 2020). There has been prominent media coverage about such risks including people with hypertension (Dunlevy, 2020; Hanrahan, 2020; Mckie, 2020), and there are concerns that people with CVD are not presenting to GPs and hospitals for management and new symptoms due to fear of contracting COVID-19 (Thornton, 2020). This study investigates whether people with hypertension have higher risk perceptions, anxiety and prevention intentions during COVID-19 restrictions, to inform targeted public health messaging for this group.

## METHODS

Data from a national Australian survey were used to conduct retrospective case-control analyses comparing hypertension and control groups. Baseline data were collected from all states/territories in April 2020 during COVID-19 lockdown, with a subsample followed-up in June 2020 when restrictions eased. Of 4362 baseline participants, those reporting high blood pressure with no other chronic conditions (n=466) were randomly matched to individual healthy controls based on age (+/-3 years), gender, education and health literacy adequacy (given baseline survey differences(McCaffery et al., 2020)). The initial match rate was 95.7%, with constraints relaxed until all cases were paired. Appropriate regression models were conducted using Stata/IC v16.1 (linear models for continuous outcomes, generalised linear models with modified Poisson approach for dichotomous outcomes, ordinal logistic regression for ordered categorical outcomes) with robust error variances to account for pair clustering, with adjustment for matching variables. A subset of the original survey cohort (n=1369) was followed-up at 2 months, including 147 participants with hypertension only, and the matching and analysis process was repeated.

The survey measures and full sample results are reported elsewhere (McCaffery et al., 2020; Wolf et al., 2020), including the Health Literacy single item screener (Wallace, Rogers, Roskos, Holiday, & Weiss, 2006), CHAI patient activation measure(Wolf et al., 2018), and State-Trait Anxiety Inventory (STAI) (Bekker, Legare, Stacey, O’Connor, & Lemyre, 2003; Marteau & Bekker, 1992). Risk perceptions and prevention behaviours were measured using Likert and categorical scales (Table 1). The perceived seriousness of the COVID-19 threat was asked generally at baseline, but at follow-up participants were asked about the public health risk generally from COVID-19, globally and in Australia specifically, given the divergent pattern of control across countries.

**Table 1.**
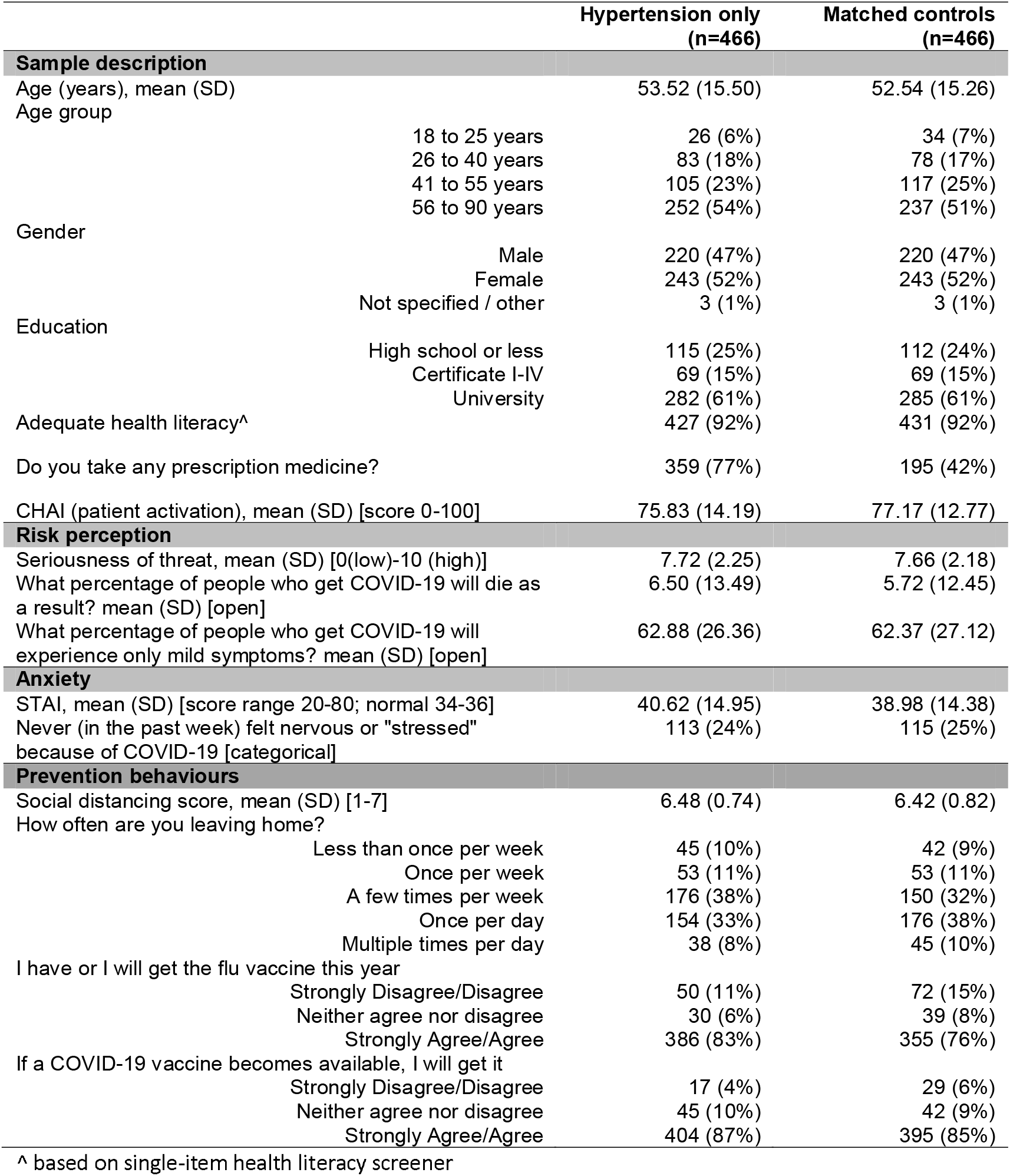
Baseline descriptive statistics and outcomes for hypertension cases versus matched healthy controls. Data are displayed as n (%) unless otherwise specified.

## RESULTS

Table 1 describes the case versus control samples for all baseline outcomes, and Table 2 shows details of the regression models comparing the two groups at this timepoint.

**Table 2.** Multivariable* regression model estimates comparing hypertension cases versus matched healthy controls at baseline.

|  | Estimate (95% CI) | p-value |
| --- | --- | --- |
| **Risk perception** |  |  |
| Seriousness of threat [Marginal mean difference; MMD] | 0.05 (-0.23, 0.34) | 0.71 |
| What percentage of people who get COVID-19 will die as a result? [MMD] | 0.75 (-0.87, 2.37) | 0.36 |
| What percentage of people who get COVID-19 will experience only mild symptoms? [MMD] | 0.71 (-2.77, 4.18) | 0.69 |
| **Anxiety** |  |  |
| STAI [MMD] | 1.90 (0.19, 3.61) | 0.03 |
| Never (in the past week) felt nervous or "stressed" because of COVID-19? [adjusted relative risk; aRR] | 0.96 (0.77, 1.19) | 0.69 |
| **Prevention behaviours** |  |  |
| Social distancing score [MDD] | 0.06 (-0.04, 0.17) | 0.21 |
| How often leaving home?^ [adjusted odds ratio [aOR] | 0.84 (0.66, 1.06) | 0.14 |
| I have or I will get the flu vaccine this year^ [aOR] | 1.52 (1.10, 2.11) | 0.01 |
| If a COVID-19 vaccine becomes available, I will get it^ [aOR] | 1.21 (0.84, 1.73) | 0.31 |

\* all multivariable models controlled for age (in years), gender, health literacy adequacy, and education; ^ analysis as ordinal logistic regression.

### Description of sample

The hypertension sample included 466 people reporting only high blood pressure and no other chronic health conditions, to isolate effects of hypertension. The average age was 53.52 years [SD 15.50], with 52% female, 47% male and 1% unspecified. The majority had a university degree (61%) and adequate health literacy (92%). The average patient activation score was comparable to other patient populations (mean scaled CHAI 74.9). Most were taking anti-hypertensive medications (77%), with 45% obtaining a refill during lockdown, 5% switching to a longer prescription, and only 1 person stopping medication.

### Risk perceptions

At baseline the perceived seriousness of threat from COVID-19 in the hypertension group was high (mean 7.72/10). On average the hypertension sample thought that 7% of people who get COVID-19 would die as a result, and 63% would only experience mild symptoms. There were no significant differences between the hypertension group and matched controls at baseline. At follow-up, those with hypertension (6.12/10) perceived a greater threat than controls (5.52/10) when asked about Australia [MMD: 0.60, 95%CI: 0.05 to 1.15; p=0.032] but not generally/globally.

### Anxiety

At baseline, 76% of the hypertension group had felt nervous or stressed about COVID-19 in the past week at least some of the time. On average the mean STAI was 1.90 units higher [95%CI: 0.19 to 3.61, p=0.03, Cohen’s d=0.13] for those with hypertension (40.75) than matched controls (38.85), with both groups higher than normal range, but below clinical levels. At follow-up there was no longer a significant difference between the hypertension (37.02) and control (36.08) groups [MMD: 0.94, 95%CI: −2.57 to 4.45; p=0.597, Cohen’s d=0.06], with anxiety reduced to within normal range in both groups.

### Prevention behaviours

At baseline the hypertension group had a social distancing score of 6.48/7 indicating strong agreement with the importance of social distancing for ones’ own health and the health of the public. Most people were leaving home a few times a week (38%) or once a day (33%) during lockdown. Overall, 83% agreed they would get the influenza vaccine and 87% would get the COVID-19 vaccine. Compared to healthy matched controls, the hypertension group was more likely to agree that they would (or have already) received the influenza vaccine this year [aOR: 1.52, 95%CI: 1.10 to 2.11, p=0.01]. There were no significant differences in willingness to vaccinate for COVID-19 (if it became available), social distancing or leaving the house. At follow-up there was no longer a significant difference between the hypertension and control groups for influenza vaccination [aOR: 1.90, 95%CI: 0.93, 3.90, p = 0.08], with intentions remaining high for both influenza and COVID-19 vaccination (>80% for both groups).

## DISCUSSION

The COVID-19 pandemic has caused major morbidity and mortality worldwide. In the current study amidst the pandemic in Australia, the main finding observed was anxiety, with higher than ‘normal’ levels during lockdown restrictions in both groups but more so for those with hypertension. This is consistent with the Australian Bureau of Statistics finding that anxiety in the general population was double the rate in April 2020 compared to a survey in 2017-18 (Australian Bureau of Statistics, 2020). Elevated anxiety during a global pandemic and unprecedented social and economic restrictions could be viewed as appropriate. On the other hand, prioritising mental health screening for more vulnerable groups with higher anxiety may be warranted given concerns about the mental health effects of COVID-19 (Wang et al., 2020). Those with hypertension were more likely to take up the influenza vaccine during lockdown compared to healthy controls. This difference in willingness does not appear to translate to a COVID-19 vaccine, but acceptance rates were high generally at baseline (Dodd et al., 2020) and even after risk perceptions and anxiety decreased when restrictions lifted.

### Strengths & limitations

The strengths of this study include a large national sample with data during and after lockdown restrictions, that enabled matched case-control analyses between participants with self-reported hypertension and healthy controls and the use of established well validated measures.

The sample was recruited via an online panel and social media, and has a low proportion of culturally and linguistically diverse participants. The survey involved non-stratified sampling without targeted recruitment of specific health conditions, and only a subset were included in the longitudinal sub-study.

### Conclusion

Anxiety was above normal levels for all groups during the COVID-19 lockdown. This was higher in the hypertension group and appeared to translate to higher influenza vaccination intentions. In Australia, where lockdown effectively reduced the spread of COVID-19 and restrictions eased relatively quickly, these differences dissipated after 2 months, but locations with prolonged restrictions may require targeted psychological screening for vulnerable groups. Despite a decrease in perceived seriousness and anxiety after 2 months of lockdown restrictions, vaccination intentions for both influenza and COVID-19 remained high (80%), which is encouraging for future prevention of COVID-19.

## Data Availability

Data available from corresponding author upon reasonable request.

## ACKNOWLEDGEMENTS

This study was not specifically funded, but in-kind support was provided by authors with research fellowships. The SHeLL group thanks the participants of the longitudinal COVID-19 survey for their ongoing participation in this research.

CB is supported by a National Health and Medical Research Council (NHMRC)/Heart Foundation Early Career Fellowship (#1122788).

CS is the recipient of a National Health and Medical Research Council (NHMRC) Practitioner Fellowship (#1154992).

RD is supported by a University of Sydney fellowship (#197589).

KM is supported by a National Health and Medical Research Council (NHMRC) Principal Research Fellowship (#1121110).

